# Associations between COVID-19 infection, tobacco smoking and nicotine use, common respiratory conditions and inhaled corticosteroids: a prospective QResearch-Case Mix Programme data linkage study January-May 2020

**DOI:** 10.1101/2020.06.05.20116624

**Authors:** Nicola Lindson, Min Gao, Jamie Hartmann-Boyce, Margaret Smith, Paul Aveyard, Duncan Young, Carol Coupland, Pui San Tan, Ashley K. Clift, David Harrison, Doug Gould, Ian D Pavord, Peter Watkinson, Julia Hippisley-Cox

**Affiliations:** University of Oxford, Nuffield Department of Primary Care Health Sciences, Radcliffe Observatory Quarter, Oxford, UK, OX2 6GG; NIHR Oxford Biomedical Research Centre, Oxford University Hospitals, NHS Foundation Trust, Oxford, UK, OX2 6GG; University of Oxford, Nuffield Department of Clinical Neurosciences, John Radcliffe Hospital, Oxford, UK, OX3 9DU; University of Nottingham, Faculty of Medicine & Health Sciences, University Park, Nottingham, UK, NG7 2RD; Intensive Care National Audit & Research Centre (icnarc), Napier House, 24 High Holborn, London, UK, WC1V 6AZ; University of Oxford, Nuffield Department of Medicine, Old Road Campus, Oxford, UK, OX3 7FZ

## Abstract

**Introduction:** Epidemiological and laboratory research seems to suggest that smoking and perhaps nicotine alone could reduce the severity of COVID-19. Likewise, there is some evidence that inhaled corticosteroids could also reduce its severity, opening the possibility that nicotine and inhaled steroids could be used as treatments.

**Methods:** In this prospective cohort study, we will link English general practice records from the QResearch database to Public Health England’s database of SARS-CoV-2 positive tests, Hospital Episode Statistics, admission to intensive care units, and death from COVID-19 to identify our outcomes: hospitalisation, ICU admission, and death due to COVID. Using Cox regression, we will perform sequential adjustment for potential confounders identified by separate directed acyclic graphs to:

1. Assess the association between smoking and COVID-19 disease severity, and how that changes on adjustment for smoking-related comorbidity.
2. More closely characterise the association between smoking and severe COVID-19 disease by assessing whether the association is modified by age (as a proxy of length of smoking), gender, ethnic group, and whether people have asthma or COPD.
3. Assess for evidence of a dose-response relation between smoking intensity and disease severity, which would help create a case for causality.
4. Examine the association between former smokers who are using NRT or are vaping and disease severity.
5. Examine whether pre-existing respiratory disease is associated with severe COVID-19 infection.
6. Assess whether the association between chronic obstructive pulmonary disease (COPD) and asthma and COVID-19 disease severity is modified by age, gender, ethnicity, and smoking status.
7. Assess whether the use of inhaled corticosteroids is associated with severity of COVID-19 disease.
8. To assess whether the association between use of inhaled corticosteroids and severity of COVID-19 disease is modified by the number of other airways medications used (as a proxy for severity of condition) and whether people have asthma or COPD.

**Conclusions:** This representative population sample will, to our knowledge, present the first comprehensive examination of the association between smoking, nicotine use without smoking, respiratory disease, and severity of COVID-19. We will undertake several sensitivity analyses to examine the potential for bias in these associations.

## Plain English summary

COVID-19 is a disease of the airways and can lead to infection of the lungs (pneumonia). Smoking is a well-known cause of airway infections. This may be because smoking makes it harder for the body to remove infectious particles or because it changes the way that the body fights infection – its immune reaction. This immune reaction causes many of the symptoms we experience when we have infections. As a result, it would seem likely that the coronavirus that causes COVID-19 could worsen infections in people who smoke. However, emerging evidence suggests that this may not be the case, and that smokers may have better outcomes than non-smokers. Some laboratory experiments have suggested that this may be due to the nicotine in cigarettes. Nicotine is available as a cheap and safe drug that could provide a treatment option for COVID-19.

People with diseases of the airways, like asthma, are at higher risk of serious lung infections. Data from China suggest that people admitted to hospital with COVID-19 are less likely to have diseases of the airways than might be expected given the rate of asthma in the general population. One reason for this could be that people with asthma or chronic obstructive pulmonary disease (COPD) take inhaled steroids as their treatment. Laboratory experiments suggest that inhaled steroids could protect against severe COVID-19.

The aim of our analysis is to assess whether smoking reduces the risk of severe COVID-19 and whether the risk of severe COVID-19 is also reduced in people who use nicotine in the form of electronic cigarettes or nicotine replacement therapy (NRT), but do not currently smoke tobacco. We also examine whether having airways disease is associated with severe COVID-19 and whether treatment for these diseases, particularly inhaled steroids, are associated with a reduced risk of serious outcomes.

We will use anonymous data from 8.3 million people’s GP records, linked to Public Health England’s database of tests for coronavirus, records of hospital admissions, records of admission to hospital intensive care units, and records of deaths due to COVID-19. We will use statistical methods to make sure we control for other factors that may cause more serious illness, resulting in admission to hospital, ICU, or death, like being older or having diabetes. Distortions in the answers we derive could result from distortions arising because of patients’ and doctors’ decision-making in who gets what kind of healthcare. We will analyse the data several ways to see if the main results are affected by these distortions that we cannot measure. These distortions can show up when the data are analysed differently.

We will be able to assess whether there is good evidence that people who smoke, use nicotine, have airway diseases, or use inhaled steroids have a different risk of experiencing serious COVID-19 when compared with the general population.

## Background rationale

### Epidemiological evidence of the association between smoking and COVID-19

Smoking is assumed to be a risk factor for COVID-19 infection and severe disease, and the WHO and other bodies have advised people to quit smoking. However, the evidence that smoking is a risk factor is mixed. A living systematic review, last updated on the 7 May 2020 found 41 studies with relevant data[1]. Of the 41 included studies, 10 explicitly recorded or reported on ‘never smoking’ status, meaning that we cannot be sure about the veracity of the coding of the remainder. Thirty-six studies appeared to show that the prevalence of smoking in hospitalised patients was lower than the national prevalence, but the quality of studies was limited (graded as fair in five studies and poor in 31 studies). Meta-analyses of two ‘fair’ quality studies suggested that current and former smokers were more likely to be tested for SARS-CoV-2, but there was no evidence of a difference in the risk of testing positive in current (RR=0.74, 95%CI = 0.31-1.73, p=0.49; I^2^ = 96%) or former smokers (RR=1.18, 95%C=0.82-1.69, p = 0.37; I^2^ = 88%) compared with never smokers. However, statistical heterogeneity was substantial and therefore these pooled estimates are unlikely to represent a true population average. Three ‘fair’ quality studies of people who tested positive for SARS-CoV-2 in the community, found no clear evidence of a decreased risk of hospitalisation among current (RR=0.95, 95%CI=0.76-1.18, p=0.62 I^2^ = 69%) or former (RR = 1.04, 95% CI = 0.98-1.10, p =0.26; I^2^ = 0%) smokers compared with never smokers. In three ‘fair’ quality studies, there was some evidence of an increased risk of greater disease severity in hospitalised current (RR=1.36, 95%CI=1.07-1.74, p = 0.01; I^2^ = 0%) but not former (RR=1.51, 95%CI=0.86-2.65, p = 0.15 I^2^ = 81%) smokers compared with never smokers. However, both statistical heterogeneity and imprecision were substantial in the latter case. A key problem with studies of hospitalised patients is that there is a high degree of selection of who is and is not admitted. Conditioning on this by confining the sample to hospitalised patients creates collider or stratification bias, that can lead to distorted associations. A good example is the obesity paradox, whereby obesity appears to be associated with a better outcomes as a result of confining analyses to hospitalised patients[2].

One problem with studies of patients in hospital is they are inevitably confounding potential effects on incidence with those on severity. We know of only one study that has thus far attempted to assess impacts on incidence. The UCL COVID-19 Social Study is a longitudinal panel survey of 53,002 adults (≥18 years) with good representation across major sociodemographic groups[3]. Compared with never smokers 0.3% [95%CI 0.2-0.3%]), the prevalence of confirmed COVID-19 was higher among current (0.6% [0.4-0.8%]) but not ex-smokers (0.2% [0.2-0.3%]). The associations were similar before (current: OR 2.14 [1.49-3.08]; ex-smokers: OR 0.73 [0.47-1.14]) and after (current: OR 1.79 [1.22-2.62]; ex-smokers: OR 0.85 [0.54-1.33]) adjustment for potential confounders. People who smoked and who had lower educational attainment were less likely to adhere to Government guidance to reduce the spread of COVID-19, and the higher risk among smokers was only evident in those with lower educational attainment, suggesting that the higher risk among smokers may be driven by behavioural factors.

A new study has much stronger data on smoking status that does not rely on self-reporting at hospital. QResearch linked 8.3 million GP records to Public Health England’s database of testing positive for COVID-19 and the ICNARC database of ICU admissions[4]. The data come from a time when the vast majority of testing for SARS-CoV-2 was in hospitalised patients with suspected COVID-19, thus swab positivity implies hospitalisation with COVID-19 in these data. The first paper from this work addresses the association between angiotensin-converting enzyme (ACE) inhibitors and angiotensin II receptor blockers (ARBs) to these outcomes. The analysis was adjusted for a wide range of confounders, including common chronic diseases, medications for these diseases, demographic and social factors, obesity, and smoking status. This showed strong evidence that smoking was associated with lower risk of hospitalisation and ICU admission. The hazard ratios (95% CIs) for hospitalisation for light, moderate, and heavy smokers were 0·49 (0·46 to 0·53), 0·39 (0·33 to 0·46), and 0·48 (0·39 to 0·59) respectively. For ICU admission, the apparent protective effect was stronger: 0·27 (0·19 to 0·36), 0·21 (0·09 to 0 ·47), and 0·11 (0·03 to 0·44) respectively. These effect sizes are so large that unmeasured confounding or bias seem unlikely explanations. A similar study using 17 million records examined the multivariable adjusted risk of COVID-19 death[5]. It reported a hazard ratio of 0.88 (0.79 to 0.99), suggesting lower protection from current smoking. A GP database study using GP-recorded smoking status reported that smokers seeking help from their GP for COVID-19 symptoms were nearly half as likely to be SARS-CoV-2 positive as never smokers 0·49 (0·34–0·71), while there was no evidence that risk in former smokers differed[6]. Taken together, the data suggest that smoking is associated with a somewhat higher risk of infection with SARS-CoV-2, but a lower risk of presenting for GP care, hospitalisation, ICU admission, and death. This suggests that smoking may reduce the severity of SARS-CoV-2 infection, but the evidence for this is uncertain.

### Mechanistic evidence on smoking and COVID-19

While it would be unthinkable to recommend smoking as treatment, nicotine is a component of tobacco, which could be used as a potential treatment. It is widely available in forms that are cheap, safe, tolerable, and non-addictive[7-9]. There are several mechanisms that could explain the effect of smoking on COVID-19, some of which relate to nicotine. First, nicotine downregulates the ACE2 receptor, to which SARS-CoV-2 attaches[10]. Second, Changeux and colleagues hypothesise that direct invasion of the olfactory nerves leading to brain invasion by SARS-CoV-2 virus is responsible for the neurological manifestations of COVID-19[11], which others have proposed plays a role in respiratory failure[12]. They point to the rabies virus which binds specifically to nicotinic receptors in the brain and hypothesise that the neurological symptoms in COVID-19 could be mediated by SARS-CoV-2 binding similarly to nicotinic cholinergic receptors, but this is highly speculative.

Nicotine also has widespread immunomodulatory effects. Nicotinic alpha-7 agonists reduce the severity of pancreatitis and peritonitis in animal models[13, 14], with infusions of nicotine causing decreased immune cell influx, pro-inflammatory cytokine release, which may be relevant in COVID-19. More generally, cholinergic receptors are widespread throughout the lung epithelium and play a role in cell division, differentiation, and apoptosis, which may be relevant to the pathogenesis of lung cancer[11].

Underlying these effects, smoking alters the distribution of T-helper cells, particularly shifting the balance between Th1, Th2, and Th17 cells, thereby influencing cytokine release, a crucial part of severe COVID-19 disease[15]. Alpha-4 and alpha-7 nicotinic receptors play important roles in regulating B cell lines[15]. Nicotine administration has also been shown to reduce the proliferation of cytotoxic T lymphocytes[16]. Nicotine has been shown to be an immunosuppressive agent that can modulate innate and adaptive immune responses[17, 18]. Stimulation of alpha-7 nicotinic acetylcholine receptors activate murine macrophages inhibiting the transcription of pro-inflammatory cytokines, which may be particularly relevant in COVID-19[19]. Furthermore, activated α7 nAChR suppressed the phosphorylation of IκB, downregulating nuclear translocation of NFκB[18, 20]. NFκB is activated in response to IL-1 and TNF-alpha release and in turn enables expression of cytokines, chemokines, and other adhesion molecules[21].

While smoking seems to be causally implicated for some diseases where the immune system plays a pivotal role, it also reduces the incidence of others including ulcerative colitis and sarcoid. Although there are many thousands of different molecules in cigarette smoke, there is strong evidence from clinical trials that nicotine alone can induce remission in ulcerative colitis,[22] providing epidemiological support to the important role of nicotine in influencing the immune response.

### Respiratory disease and COVID-19

In the COVID-19 pandemic, attention has focused on cardiovascular diseases, but pre-existing respiratory disease is potentially relevant in determining the severity of COVID-19 pneumonia, the main cause of adverse outcomes. A review of case series showed that the prevalence of chronic respiratory disease in patients hospitalised with COVID-19 was lower than its prevalence in the general population[23]. As with smoking, one possible explanation is that respiratory disease was unrecorded on admission, but the same series showed that diabetes was more commonly recorded than its background prevalence, which perhaps suggests this is an unlikely explanation. Another explanation is that inhaled corticosteroids (ICS) may modify the severity of COVID-19. Studies show reduced coronavirus replication and cytokine production in cell lines treated with steroids and beta agonists[24, 25]. In contrast, there is observational evidence and evidence from randomised trials that ICS is associated with an increased risk of non-COVID-19 upper and lower respiratory tract infections[26, 27]. Alternatively, the association between respiratory disease and may be a selection bias since asthma and COPD were identified as high-risk conditions during the pandemic with those affected advised to take precautions such as self-isolation. Finally, people who smoke are more likely to be using inhalers for COPD or asthma and ICS use could explain the unexpected findings on smoking or vice versa.

A systematic review aimed to assess the association between prior treatment with ICS and COVID-19 severity[26]. After screening 771 publications, 59 were identified for full text screening. No studies were identified that examined the association between prior treatment with ICS and outcomes in COVID-19.

### Summary of background and aims

Smoking could be associated with a modest increased incidence of COVID-19 disease, which may be related to behavioural factors. However, there is now some evidence that the incidence of more severe COVID-19 may be lower in people who smoke and, in particular, that the risk of ICU admission may be lower, with some suggestion of a dose-response effect. Smoking involves inhaling several thousand different molecules and has widespread immunomodulatory actions. However, it is clear from experimental work that nicotine alone has important immunomodulatory actions (generally suppressing inflammatory responses), and can put ulcerative colitis into remission. Nicotine in the form of nicotine replacement therapy (NRT) poses few risks of addiction in those not already addicted to it, is widely available, cheap, safe, and tolerable for most users, including non-smokers, and could represent a feasible treatment for COVID-19. Likewise, there appears to be a reduced incidence of hospitalisation in people with chronic respiratory conditions, against expectations, and mechanistic evidence that inhaled corticosteroids could be one explanation. Given the overlap between respiratory disease and smoking, we examine also whether prior use of ICS is associated with severity.

The aims of this analysis are:

1. To assess the association between smoking and COVID-19 disease severity, and how that changes on adjustment for smoking-related comorbidity.
2. More closely characterise the association between smoking and severe COVID-19 disease by assessing whether the association is modified by age (as a proxy of length of smoking), gender, ethnic group, and whether or not people have asthma or COPD.
3. To formally assess for evidence of a dose-response relation between smoking intensity and disease severity, which would help create a case for causality.
4. To examine the association between former smokers who are using NRT or are vaping and disease severity.
5. To examine whether pre-existing respiratory disease is associated with severe COVID-19 infection.
6. To assess whether the association between chronic obstructive pulmonary disease (COPD) and asthma and COVID-19 disease severity is modified by age, gender, ethnicity, and smoking status.
7. To assess whether the use of inhaled corticosteroids is associated with severity of COVID-19 disease.
8. To assess whether the association between use of inhaled corticosteroids and severity of COVID-19 disease is modified by the number of other airways medications used (as a proxy for severity of condition) and whether people have asthma or COPD.

## Methods

### Design

This is a prospective cohort study of patients registered in English general practices linked to data from Public Health England’s (PHE) database of SARS-CoV-2 positive tests, to Hospital Episode Statistics (HES), to data on patients admitted to all English intensive care units (ICU) provided by linking to Intensive Care Research and Audit Centre (ICNARC), and to the Office for National Statistics (ONS) data on death certificates. This protocol is written following RECORD guidance[28]. These linkages allow us to assess the association between our exposures and severe COVID-19.

### Setting

1205 general practices in England contributing to QResearch.

### Participants

People registered as patients and aged 20-99 years on 1 Jan 2020 who were registered for the whole of 2019 and remained so until censoring at 10 May 2020 for linkage to PHE SARS-CoV-2 data, at 31 March 2020 for HES data, and 13 May 2020 for linkage to ICNARC data, and 28 April 2020 for linkage to ONS data.

### Outcomes

We have three outcomes, and we will interpret the data based on the associations seen for all three outcomes, looking for consistency of association and interpretable patterns. The outcomes are:

1. Death from COVID-19. This is defined as (a) cause of death certificate (ICD10-codes for confirmed or suspected COVID) or death in someone who has tested positive for COVID-19 infection.
2. Hospital admission for COVID-19 defined as testing positive for SARS-CoV-2 and appearing in the HES dataset as an in-patient.
3. Admission to ICU with severe COVID-19.

### Exposures

1. For the analysis of smoking, we will code smoking as: never smoking, ex-smoker, light (1-9 cigarettes/day), moderate (10-19 cigarettes /day) or heavy smoker (20+ cigarettes per day) as this is recorded in the GP records. We will use the most recent recorded value of smoking status at the study start date.
2. For the analysis of NRT, we will code a person as an NRT user if the last prescription for NRT was in the month prior to testing SARS-CoV-2 positive or admission to ICU with COVID-19.
3. For the analysis of vaping, we will classify someone as vaping if they have a code indicating that since cohort inception and no subsequent code indicating that they have stopped vaping.
4. For the analysis of respiratory disease (code group ID), we will define the diseases of interest as COPD (29), asthma (28), bronchiectasis (1829). If there are sufficient numbers of patients, we will also examine associations between COVID-19 severity and cystic fibrosis (303), sarcoidosis (6501), extrinsic allergic alveolitis/hypersensitivity pneumonitis (6503), idiopathic pulmonary fibrosis (6502), other interstitial lung disease (6504), and lung cancer (51).
5. For the analysis of ICS, we will code prior ICS use if they had a prescription for ICS in the two months prior to SARS-CoV-2 positivity or ICU admission.

### Potential confounders

Our approach to confounding is guided through directed acyclic graphs (DAG) to identify potential confounders, colliders, and biasing relationships.

**Figure 1.**
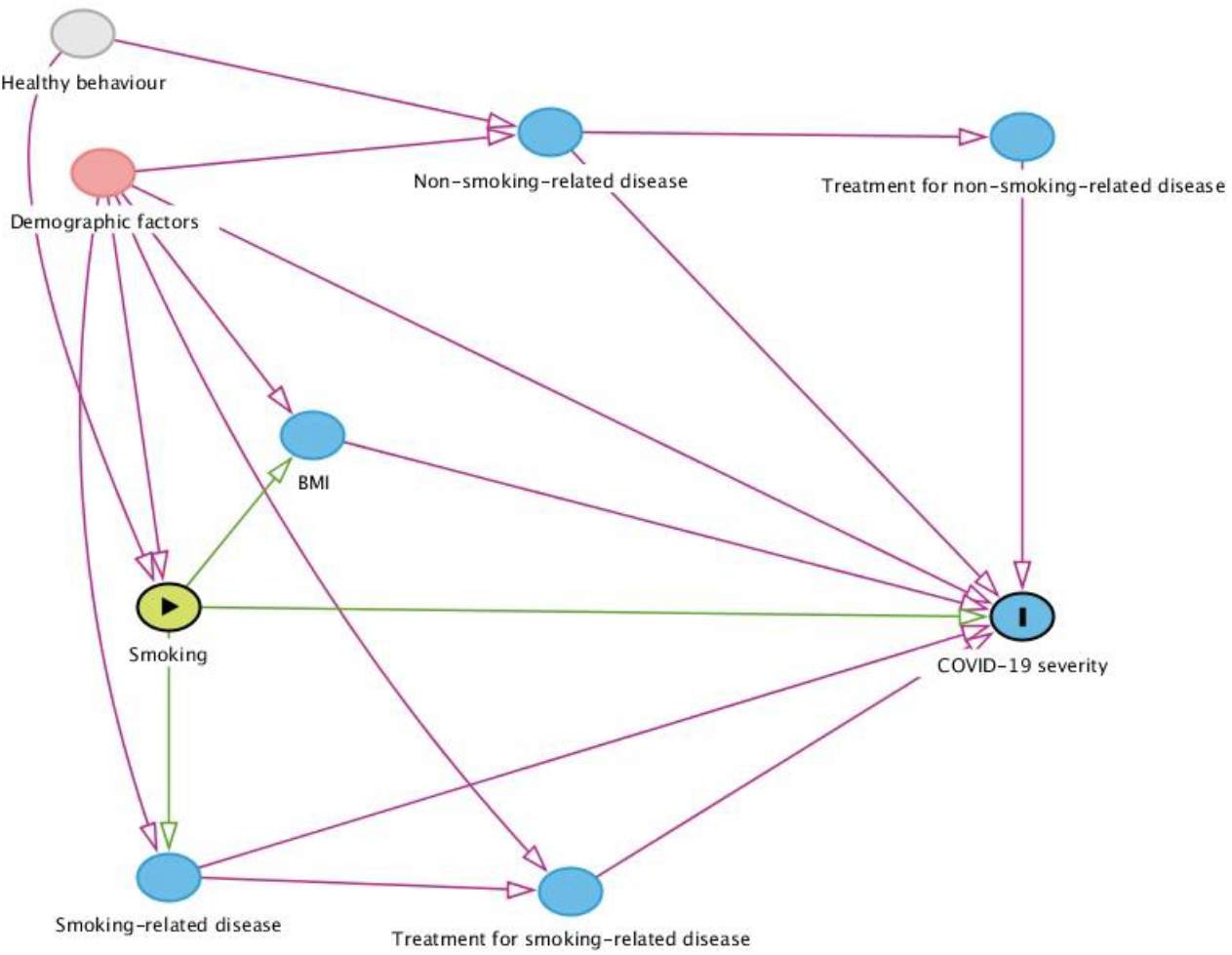
DAG for the association between smoking and severe COVID-19. In this and subsequent DAGs, green is the exposure variable, and the blue variable with the I is the outcome. Causal paths are shown in green, while biasing paths are shown in red.

This DAG shows four paths between the exposure and the outcome- the direct path, the path via BMI, the path via smoking-related disease, and the path via treatment for smoking-related disease. There are two back door (confounding) paths- via demographic factors and via non-smoking-related disease. Thus, the minimally sufficient adjustment to get the total effect of smoking on COVID-19 outcomes is to adjust for demographic factors and non-smoking-related diseases. (The path through treatment for non-smoking-related disease is blocked by controlling for non-smoking-related disease). To isolate the direct effect, the effect of interest here, we must therefore also adjust for the three mediating paths via BMI, smoking-related disease, and treatment for smoking-related disease.

**Figure 2.**
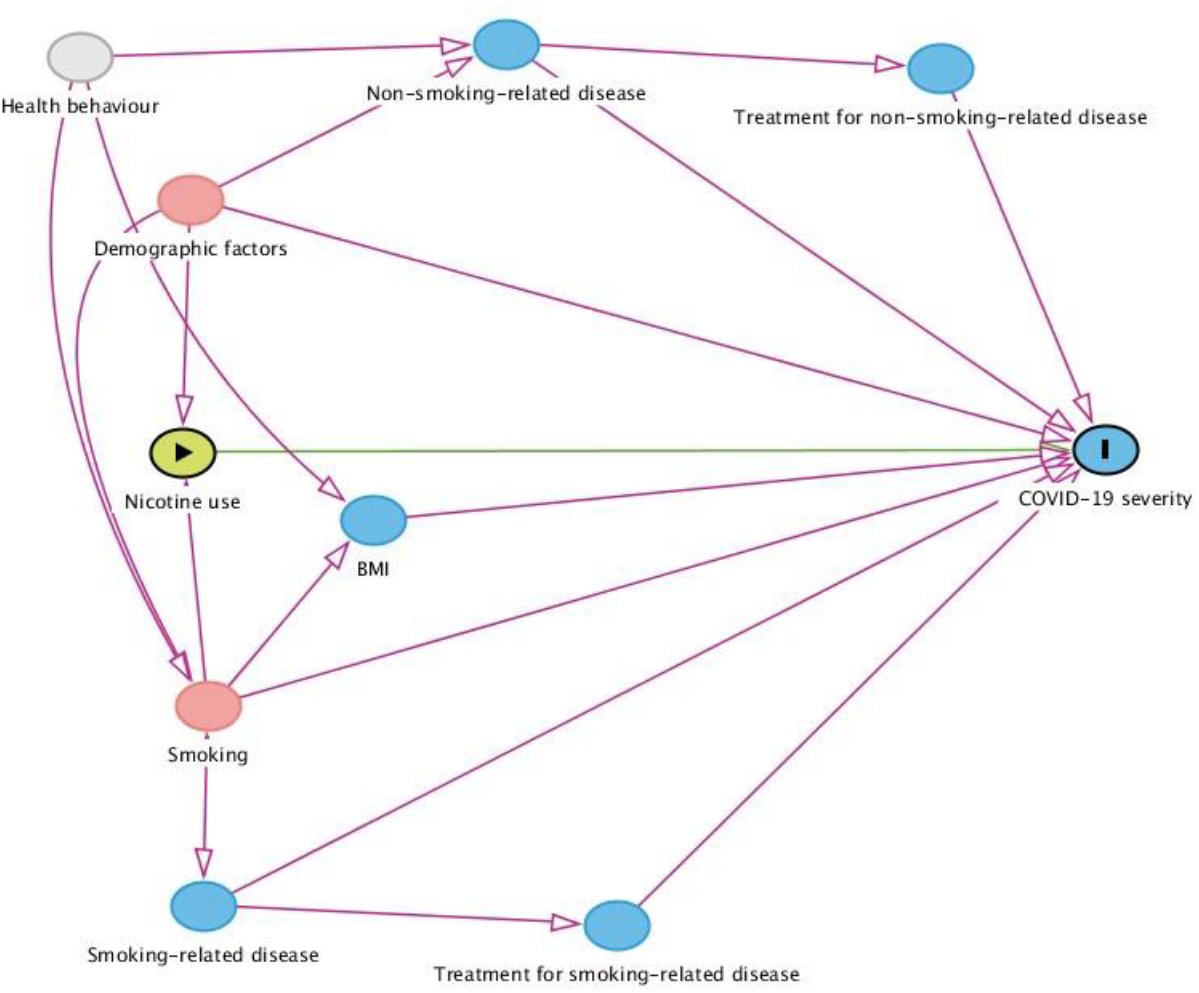
DAG for association between nicotine use and severe COVID-19. Nicotine use here means either NRT use or vaping.

This model greatly simplifies the adjustment set. There is one causal path between nicotine use and the outcome. There are two back door, potentially confounding paths between nicotine use and COVID-19 severity, one via demographic factors and one via smoking. Controlling for these two factors completely blocks all other backdoor paths and thus provides a minimally sufficient set of controls to estimate the total effect, which is equal to the direct effect in this case.

**Figure 3.**
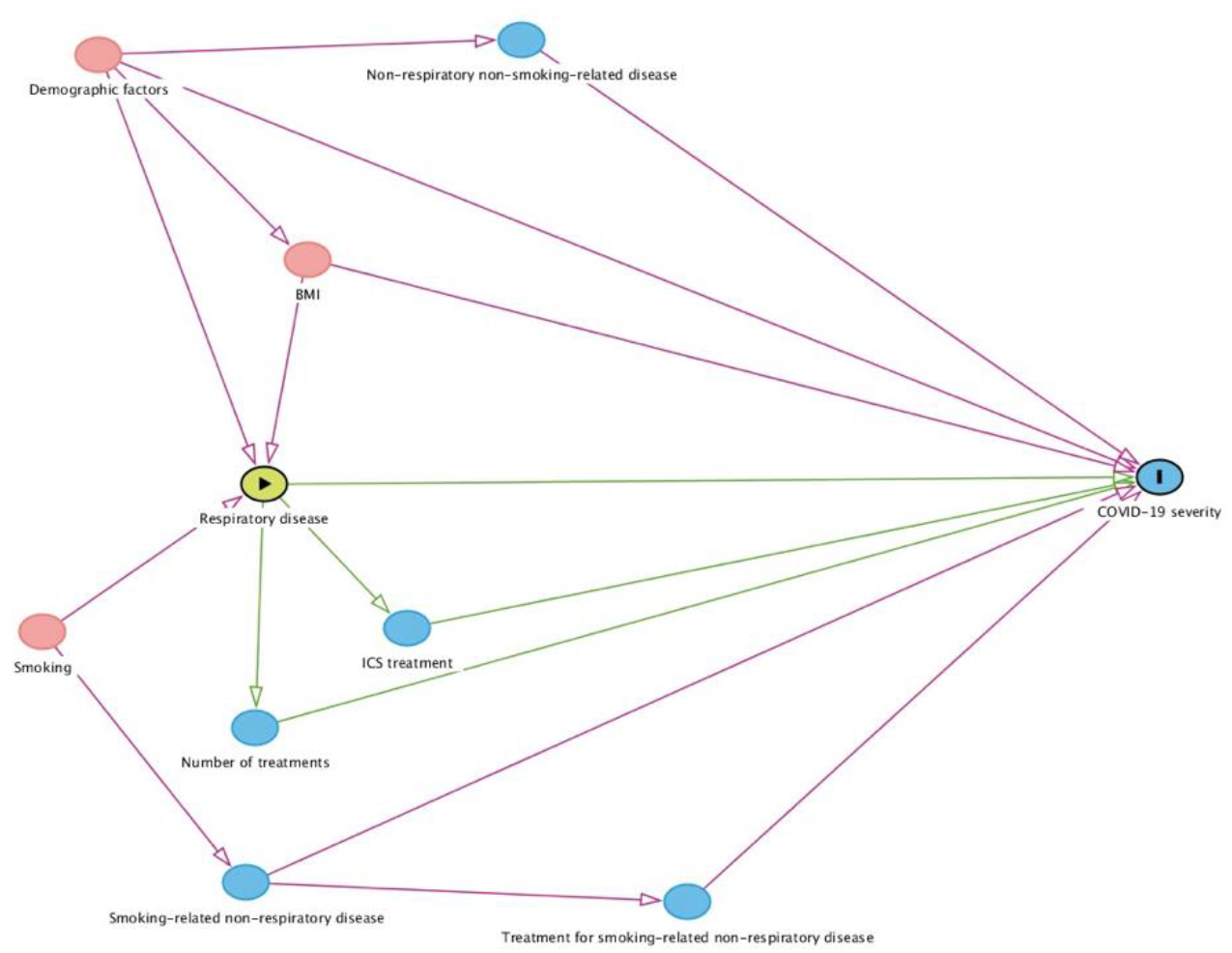
DAG for association between respiratory disease and severe COVID-19.

This DAG shows three paths for the total effect of respiratory disease on COVID-19 severity: direct, via ICS, and via number of treatments. There are three back door paths for potential confounding that are completely blocked by controlling for smoking, BMI, and demographic factors. (Healthy behaviour, a latent variable in this dataset, would create paths between smoking, BMI and non-smoking-related disease that would not create any further backdoor paths and thus would be controlled by the existing set). To isolate the direct path between respiratory disease and outcome would require us to also control for ICS treatment, and number of treatments.

This DAG shows one direct path, which is the only causal path between ICS use and severe COVID-19. The only back door and therefore potentially confounding path is by respiratory disease, so controlling for this single variable will block all other paths to the outcome.

The DAGs are drawn conceptually to represent slightly different collections of variables. The variables that are common to all the DAGs are

1. Demographic variables age, sex, ethnicity (White, Indian, Pakistani, Bangladeshi, other Asian, Black African, Chinese, and other ethnic group), material deprivation by quintile of Townsend score, and geographical region of England.
2. Smoking-related morbidity. This is divided into respiratory disease- COPD, asthma, lung cancer, and idiopathic pulmonary fibrosis, and smoking-related non-respiratory disease coronary heart disease, stroke, heart failure, atrial fibrillation, type 2 diabetes, and chronic kidney disease defined as eGFR<60ml/minute/1.73m^2^.
3. Non-smoking-related morbidity divided into respiratory disease- bronchiectasis, cystic fibrosis, sarcoidosis, extrinsic allergic alveolitis/hypersensitivity, and other interstial lung diseases; and non-respiratory- type 1 diabetes, chronic liver disease, such as hepatitis, chronic neurological conditions, such as Parkinson’s disease, motor neurone disease, multiple sclerosis (MS), a learning disability, cerebral palsy, and (not a disease) pregnancy.
4. Treatment for smoking-related morbidity- common diseases for cardiovascular disease-statins, ACE inhibitors, angiotensin 2 receptor blockers, beta blockers, calcium channel antagonists, thiazides, potassium sparing diuretics, anticoagulants, antiplatelet agents, treatments for type 2 diabetes- biguanides, sulphonylureas, other diabetes drugs, ICS, number of other agents for airways disease.
5. Number of medications used for airways disease by class (beta adrenergic agonists, muscarinic agonists, leukotriene receptor antagonists, and xanthines).
6. Body mass index (classified as<18.5; 18.5-24.99, 25-29.99, 30-34.99, 40+ kg/m^2^).

**Figure 4.**
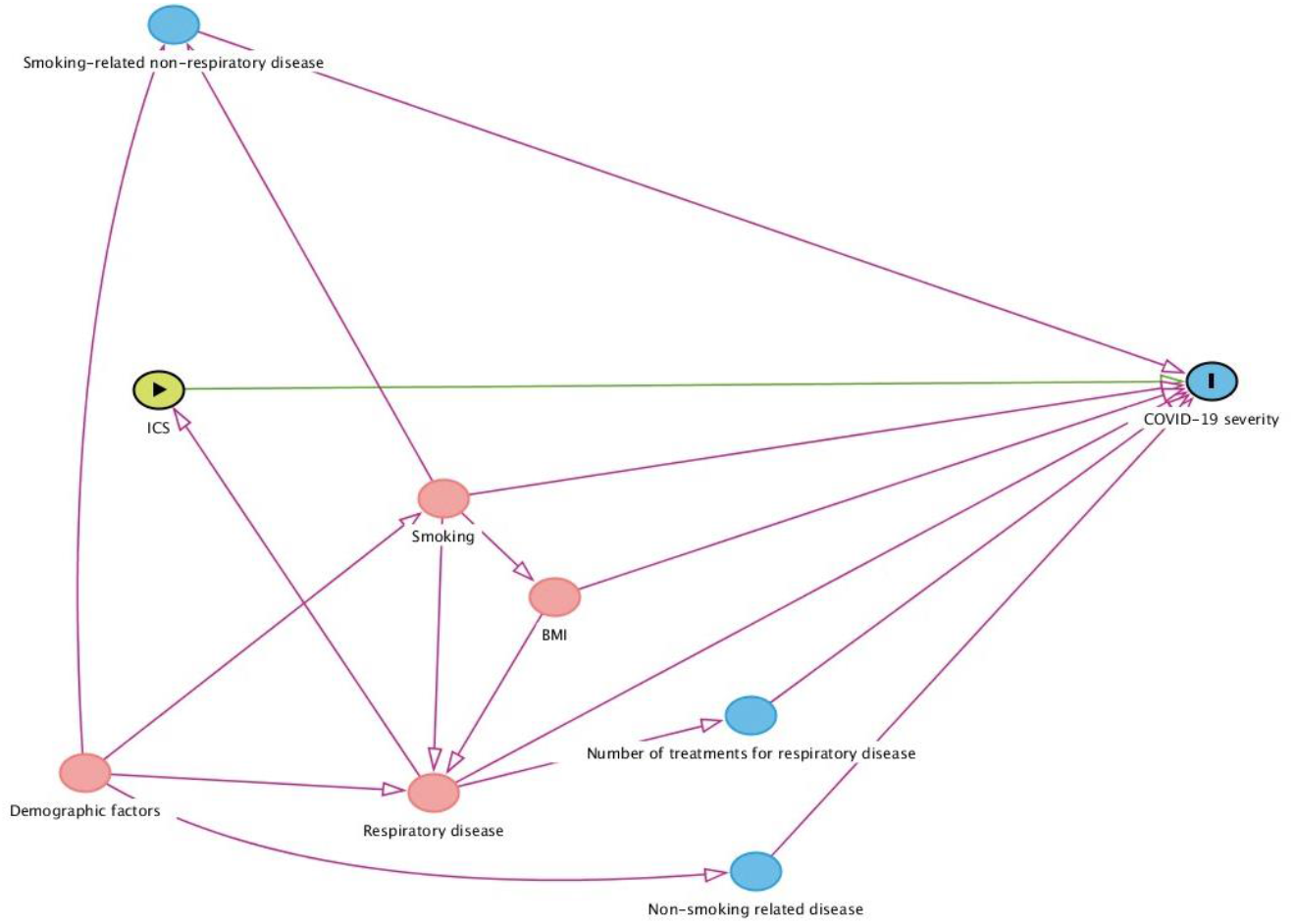
DAG for the association between ICS use and COVID-19 severity.

### Effect modifiers

For the association between smoking and severe COVID-19 disease, we will assess whether the association is modified by age (as a proxy of length of smoking), gender, ethnic group, and whether or not people have asthma or COPD by including a multiplicative interaction term and reporting appropriate p values. This analysis is exploratory.

For the association between COPD and asthma and severe COVID-19, we will assess whether this is modified by age, gender, ethnicity, and smoking status by including a multiplicative interaction term and reporting appropriate p values. This analysis is exploratory.

For the association between ICS use and severe COVID-19, we will assess whether this is modified by the number of other airways medications used (as a proxy for severity of condition) and whether people have asthma or COPD by including a multiplicative interaction term and reporting appropriate p values. This analysis is exploratory.

### Data sources

The outcome data come from three sources, the national intensive care Case Mix Programme (covering all of England) and Public Health England SARS-CoV-2 database of positive tests; HES and ONS civil registration (mortality) data. All data other than outcome data comes from QResearch.

### Bias

Smoking status of patients in GP records closely matches national prevalence data[29]. However, smoking status is incomplete for a small minority of people in GP records. In the main analysis we will impute these data (see below), but in sensitivity analysis confine the analysis to people with recorded data. Smoking status may be out of date because people who have given up have not been recorded as such or some who were recorded as former smokers may have relapsed. This is likely to result in non-differential misclassification generally reducing the strength of association[30]. We will address this in sensitivity analysis by confining the analysis to people where smoking status has been recorded in the year prior to the index outcome.

It is plausible that people who smoke may choose not to present for healthcare or their clinicians diagnose their condition as something other than COVID-19 or avoid referral to hospital on the grounds of frailty associated with smoking. This would cause a spurious association between smoking and reduced severity of COVID-19. We will address this by sensitivity analysis, excluding from the analysis people who smoke and have other comorbid conditions, which may be most likely to be misdiagnosed or not offered care in an intensive care unit.

To assess the extent of possible unmeasured confounding, for the hazard ratios derived from Cox regression models, we will derive the “E-value”[31]. This is defined as the minimum strength of association that an unmeasured confounder would need to have on both exposure and outcome (conditional on measured covariates) to adjust away any observed exposure-outcome association. E-values therefore represent a measure of the robustness of any association, and large values suggest that large unmeasured confounding is required to ‘explain away’ any observed effect.

The standard approach to most analyses is to weight each observation individually. However, in some cases, the probability of exposure may also be affected by factors that also affect the outcome. As some drivers of the exposure also are also possibly linked to outcomes, this may represent a selection bias[32]. Inverse probability weighting (IPW) is one method that seeks to mitigate such selection bias. Here, the ‘weights’ assigned to each observation are based on each person’s probability of being exposed given the presence of confounders. As one sensitivity analysis, we will utilise IPW methods to estimate the ‘treatment effect’ of smoking status on our primary outcome. We will perform logistic regression analyses to estimate the likelihood of being a smoker based on demographic factors and comorbidites and use normalised weights therefrom to adjust the estimation of the effects of smoking on death from COVID-19. Therefore, we will observe whether the unweighted and weighted analyses are similar in direction and magnitude of association.

### Study size

To maximise precision and minimise selection bias we will include all patients registered within the QResearch database on (01.01.2020). To detect a HR of ≥1.2 with 90% power, 1% significance, for an exposure with 20% exposed then 2515 outcome events are needed. To detect a HR of ≤0.8 with 90% power, 1% significance, for an exposure with 10% exposed then 2143 outcome events are needed. For smoking, with a prevalence of 17% exposed, then to detect HR≥1.2 then 2819 events are needed, and for HR≤0.8 then 2461 outcome events are needed.

### Quantitative variables

Most variables in this analysis are categorical and will be analysed using the natural reference category as the reference group. Continuous variables like BMI will be categorised to speed the analysis on this large data set. It will be categorised as described above as these are WHO categories.

### Statistical methods

GP data on smoking status, body mass index, and ethnicity is incomplete. To allow data analysis with all participants, we will impute missing data. After conducting univariable analyses, we will conduct a multivariable analysis with complete data to predict smoking status, body mass index and ethnicity. We will then use multiple imputation with chained equations to replace missing values for smoking status, body mass index and ethnicity and use these values in our main analyses[33]. We will include all exposure and explanatory variables in the imputation model, along with the Nelson–Aalen estimator of the baseline cumulative hazard, and the outcome indicator. We will carry out five imputations.

For all analyses, we will use Cox proportional hazard models, with follow up time as the timescale. We will use p< 0·01 (2-tailed) to determine statistical significance. We describe below the modelling strategy. Taking account of potential confounders also opens the possibility of biasing pathways, so we will take note of whether adjustments lead to unexpected shifts in coefficients of the variable of interest and that might indicate this.

1. To assess the association between smoking and disease severity and how that changes on adjustment for smoking-related comorbidity. We will first conduct univariable analysis for the association between smoking as categorised above on COVID-19 outcomes. We will then sequentially adjust using our DAG as a guide. We will estimate then adjust for demographic factors and non-smoking-related disease. To isolate the direct effect, the effect of interest here, we must therefore also adjust for the three mediating paths via BMI, smoking-related disease, and treatment for smoking-related disease.
2. More closely characterise the association between smoking and severe COVID-19 disease by assessing whether the association is modified by age (as a proxy of length of smoking), gender, ethnic group, and whether people have asthma or COPD. Using the final model described in 1, above, as the base model we will add appropriate multiplicative interaction terms for the effect modifiers. These will be added one at a time and removed before testing for further interactions. Where there is evidence of more than one interaction, we will add all such to one model.
3. To formally assess for evidence of a dose-response relation between smoking intensity and disease severity. Using the final model described in 1, above, as the base model we will replace the categorical variables for current smoking with a linear variable coded 1, 2, and 3 to represent the categories of intake in a single variable. The exact daily consumption of cigarettes is unknown, but this scale is approximately proportional to the median category values of cigarettes/day (5, 15, 25) as coded in QResearch. We will assess the association between smoking coded in a dose-response manner and our outcomes.
4. To examine the association between former smokers who are using nicotine replacement therapy or are vaping and disease severity. This analysis will be confined to people coded as either current or former smokers. Given that GPs are likely to have very variable practices at recording quit attempts, we will assume that everyone prescribed NRT in the past month is attempting to quit. Using the DAG in Figure 2, we will estimate the unadjusted association between nicotine use indicated by vaping or NRT use and our outcomes. We will then control potential confounding by controlling for smoking status and demographic factors to provide our best estimate of the unconfounded effect.
5. To examine whether pre-existing respiratory disease is associated with severe COVID-19. We will examine the unadjusted association between each individual respiratory disease outcomes. We will then examine the total effect of respiratory disease on COVID-19 severity by adjusting for smoking, BMI, and demographic factors. This is the association of most interest, but we will examine the direct effect only by controlling for ICS use and number of treatments.
6. To assess whether the association between COPD and asthma and COVID-19 disease severity is modified by age, gender, ethnicity, and smoking status. For the multivariable model described in 5 for the total effect, we will add multiplicative interaction terms individually, and testing for statistical significance of the interactions across strata using likelihood ratio tests.
7. To assess whether the use of inhaled corticosteroids is associated with severity of COVID-19 disease. Figure 4 shows one direct path, which is the only causal path between ICS use and severe COVID-19, for which we will estimate the size of the association. We will then adjust for the only necessary potential confounder, which is the presence of coded respiratory disease. Given that most people coded with asthma in the GP database are likely to be using ICS and many with COPD, this could introduce unacceptable collinearity. If so, we will remove the terms for asthma and COPD. If so, it is not possible to block all confounding paths. However, controlling for number of treatments for respiratory disease, BMI, smoking, and demographic factors would provide partial control that is unlikely to cause collinearity (Figure 4).
8. To assess whether the association between use of inhaled corticosteroids and severity of COVID-19 is modified by the number of other airways medications used (as a proxy for severity of condition) and whether people have asthma or COPD.

We will add appropriate multiplicative interaction terms to the model described in 7, individually, then together if there are more than one significant interaction.

### Sensitivity analyses

Some sensitivity analyses are described in the risk of bias section. For the analysis between ICS use and severity, respiratory disease is coded only in a binary present/absent fashion. It is not possible to quantify severity of disease. Instead, we could conceptualise a latent variable, disease severity, which would have causal links from it to ICS use and to number of treatments for respiratory disease. This would open a potential confounding path, which would be partly blocked by controlling for number of treatments.

We recognise that relatively few former smokers will be coding as vaping or have recently used NRT in the database and that analyses on our three outcomes may lack precision. If so, we will therefore create a composite outcome that comprises admission to hospital with COVID-19 or death from COVID-19.

### Sub-group analyses

If there is evidence of significant interactions, we will assess the strength of associations in separate subgroups for clarity of presentation.

### Data access and cleaning methods

Person-level data linkage will be undertaken between QResearch, Hospital Episode Statistics, the Case Mix Programme and Public Health England. Cases will be matched on pseudonymised NHS number[34].

The authors will have full access to anonymised data from the Case Mix Programme, QResearch and Public Health England, and Hospital Episode statistics databases. The following data cleaning will be undertaken on these databases: duplicates will be removed, dates and values outside a credible range were set as missing.

## Discussion

This protocol describes what will be by far the largest linkage of primary and critical care data to investigate the associations between smoking, nicotine replacement, chronic respiratory disease (and drugs used to treat it) and severe COVID-19 disease.

We will use four high quality, established large validated research databases (QResearch and ICNARC CMP, HES and ONS) and link them to the national register of SARS-CoV-2 test results. Our study is observational with strengths and inherent limitations since the data were collected as part of routine NHS care. Key strengths include the use of high quality, established validated databases, size, representativeness, lack of selection, recall and respondent bias. UK general practices have good levels of accuracy and completeness in recording clinical diagnoses and prescriptions and provide the ability to update analyses as data changes over time[35]. It is therefore likely to be representative of the population of England. It has good face validity since it has been conducted in the setting where most patients in the UK are assessed, treated and followed up.

It is likely that prescribed NRT will under-ascertain exposure to nicotine replacement therapies because most NRT is purchased not prescribed and this may particularly have been the case during the early stages of the COVID-19 pandemic. However, this does not invalidate the analysis because it is likely that the majority of people prescribed NRT will be using it and therefore the association will be largely unbiased. We cannot exclude the possibility that smoking history is related to being kept at home and/or not given ICU care because of the degree of underlying chronic disease and that this will affect our findings. However, we will explore this possibility by examining the presence of smoking-related co-morbidities in the primary care and hospital COVID positive populations. There may be some over-ascertainment of exposure to medication since our definition is based on issued prescriptions rather than dispensed medication. Data on community and care home deaths or deaths occurring within hospital but not in ICU are not yet available from Hospital Episode Statistics and Civil Registrations. Linkage of the GP data to national registries of outcome data, updated in near real time, will have minimised ascertainment bias relating to laboratory confirmed cases. However, there will be under-ascertainment of total COVID-19 cases due to the current absence of widespread systematic testing strategy in the UK, and due to false negative tests. As UK health policy during the study period confined testing for COVID-19 almost entirely to hospitalised patients, our data focuses on the incidence of more severe disease, rather than all cases, as most people with probable COVID-19 are not admitted to hospital. Not all acutely unwell patients who further deteriorate in hospital are admitted to ICU and this may result in bias. Admission to ICU is limited to those who might benefit from this treatment and so varies on the basis of patient demographic and medical characteristics. Data on deaths on ICU were available to us but a significant proportion of patients admitted to an ICU were still being treated in an ICU and this varied by region as the pandemic spread. For this reason, ICU deaths were not included in the analysis.

## Data Availability

Not applicable

